# Predicting a metachronous cutaneous squamous cell carcinoma: a competing-risk model based on nationwide linked registries

**DOI:** 10.64898/2025.12.18.25342538

**Authors:** Andrya Reder Hollatz, Celeste J. Eggermont, Barbara Rentroia-Pacheco, Marieke Louwman, Antien Mooyaart, Tamar Nijsten, Marlies Wakkee, Loes Hollestein

## Abstract

**Background:** following a first cutaneous squamous cell carcinoma (CSCC), one-third of patients develop new primaries, escalating their risk of metastasis and poor outcomes. However, current follow-up strategies are not risk-stratified, representing a critical gap in patient management.

**Objective:** to develop and validate a prognostic model to quantify individualized absolute risk of a first metachronous CSCC after an index tumor, accurately accounting for the high competing risk of mortality in this typically elderly population.

**Methods:** we conducted a nationwide, population-based cohort study of 11,737 patients with a first histologically confirmed CSCC (Netherlands Cancer Registry, 2007–2008) with up to 10 years of follow-up. Data on subsequent tumors was retrieved via linkage to the Automated National Pathological Anatomy Archive (Palga). A Fine–Gray competing-risk model was developed using routinely available clinical and pathological predictors (age, sex, hematologic malignancy, basal cell carcinoma (BCC) and actinic keratosis (AK) history, presence of synchronous CSCC, primary tumor location, and differentiation). Model performance was assessed 10-fold cross-validation, quantifying discrimination (time-dependent C-index) and calibration.

**Results:** during follow-up, 3,288 (28%) developed a first metachronous CSCC. The model identified key predictors: markers of cumulative UV-exposure (included AK history, ≥5 prior BCCs), and immunosuppression (chronic lymphocytic leukaemia/small lymphocytic leukaemia). Male sex, presence of synchronous CSCC at baseline were also associated with higher risk. While discrimination was modest (cross-validated 5-year C-index: 0.64), the model demonstrated excellent calibration.

**Conclusions:** this competing-risk model provides individualized, well-calibrated absolute risk estimates for a first metachronous CSCC. Based on routinely available clinical features, it offers insight into how established predictors shape risk in this high-susceptibility population. External validation and the identification of novel predictors are necessary to further refine the model and support personalized dermatologic care.

## INTRODUCTION

Cutaneous squamous cell carcinoma (CSCC) is the second most common skin cancer, with incidence rates continuing to rise globally. ^1,2^ Following a first CSCC, approximately one-third of patients develop at least one metachronous (i.e., independent primary that develops after a first tumor) CSCC.^3–5^ Consequently, due to tumor multiplicity, the true burden of CSCC on healthcare systems exceeds simple incidence rates by approximately 1.5-fold. ^6–8^ Additionally, although follow-up frequency varies in clinical practice, European and American consensus guidelines still recommend routine long-term surveillance, with at least annual visits for patients after a first CSCC, further increasing healthcare load.^9–12^

Patients with multiple CSCCs are at higher risk for poor outcomes,^13^ and the recently developed Erasmus MC model identified the number of previous CSCCs as a predictor for metastasis.^14^ Because it remains difficult to predict which patients will develop another tumor, this routine long-term surveillance focus is currently clinically justified. However, the risk of developing metachronous tumors varies considerably among patients, with more than half of them remaining disease-free after 10 years ^15–17^

Risk factors for multiple CSCCs are described, such as history of non-melanoma skin cancer, immunosuppression, advanced age and cumulative sun exposure.^18–20^ Yet, few studies focused on individual risk estimates and profiling for a metachronous CSCCs. ^21–23^ This lack of risk stratification makes universal long-term follow-up increasingly unsustainable, resulting in over-surveillance of low-risk patients and, potentially, insufficient monitoring of high-risk individuals; there is thus a critical need to identify which patients are at greatest risk for metachronous CSCCs, as well as candidates for de-escalated surveillance.

We therefore sought to develop a prediction model to estimate the absolute risk of a first metachronous CSCC after a primary tumor, using data from a nationwide cohort of >12,000 patients with long-term follow-up from the Netherlands Cancer Registry (NCR).

## METHODS

### Study Design and cohort definition

The study population consists of an updated cohort previously described. ^24^ In short, all patients with a first, histologically confirmed primary CSCC in 2007 or 2008 were identified from the NCR. Patients were followed from the date of first CSCC until death or end of linkage period (August 26, 2020), whichever occurred first. Vital status was retrieved from municipal records updated through January 31, 2022.

As tumors after a first primary CSCC were not registered by the NCR up to mid-2016, we identified new follow-up records via linkage to the Automated National Pathological Anatomy Archive (Palga).^25^ To ensure correct coding as new primary CSCCs instead of recurrences, we applied a rule-based algorithm using International Classification of Diseases for Oncology (ICD-O-3) anatomical subsites and same lateralization combined with a 3-month time window.^26^ Histologically confirmed diagnoses of basal cell carcinoma (BCC) and actinic keratosis (AK) occurring before the index CSCC were also retrieved via Palga linkage. The study protocol was approved by the scientific and privacy committees of the NCR and Palga.

Solid organ transplant recipients (SOTR) were excluded from the cohort, since their distinct and exceptionally high risk of multiple CSCCs introduce significant bias into a general population model. Additionally, follow-up guidelines recommending intensive follow-up already exist for this subgroup.^27–30^ The development cohort comprised then 12,122 unique patients (Figure 1).

**Figure 1.**
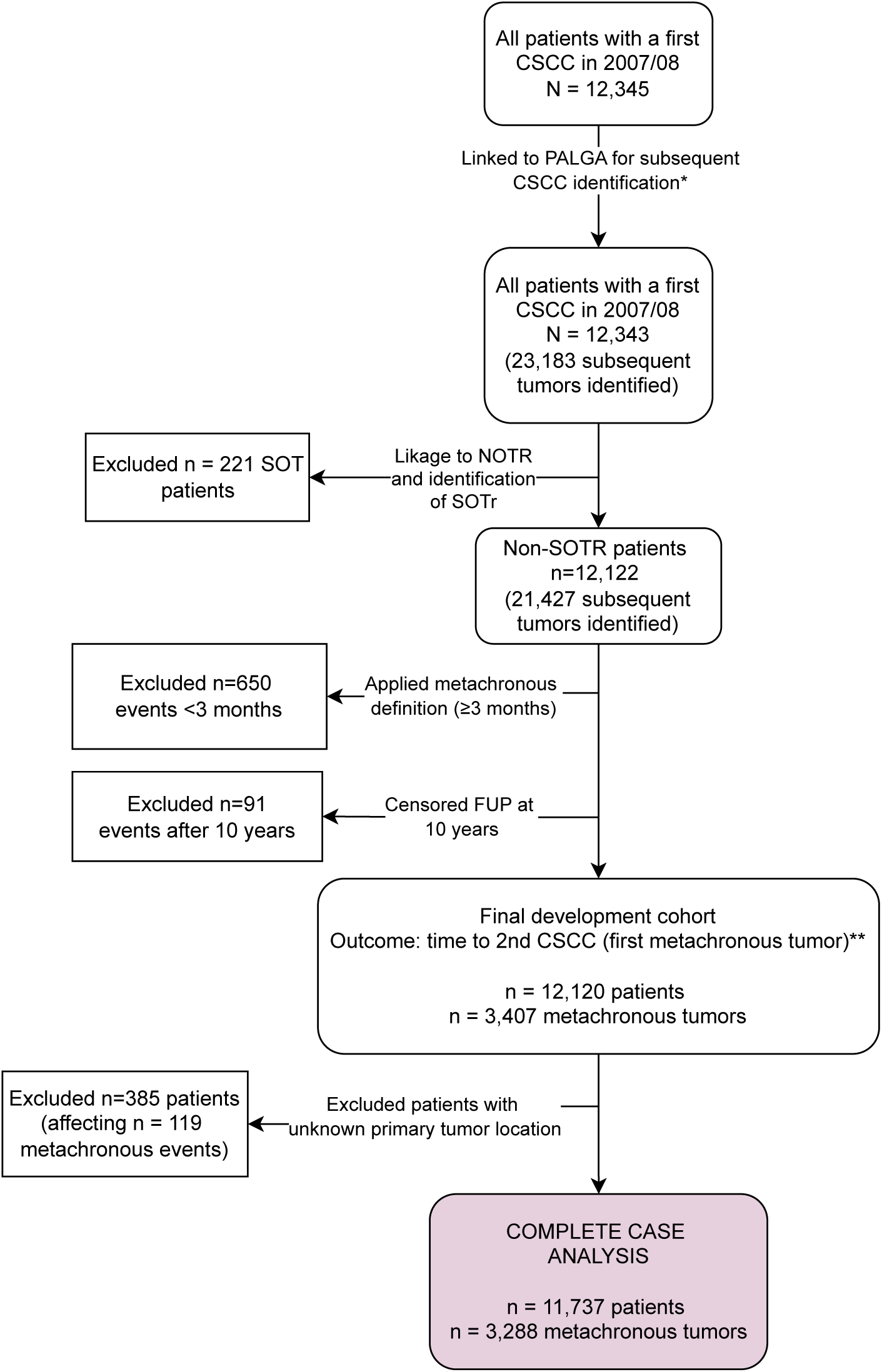
Flowchart of patient selection for model development. From N=12,345 patients with a first CSCC in 2007–2008, 11,737 remained in the final model after exclusions. *126 suspected recurrences excluded; 2 vulvar cases removed after review. **Time to 3rd–4th CSCCs not modeled (n=1750, 940). Abbreviations: CSCC, cutaneous squamous cell carcinoma; NOTR, Netherlands Organ Transplant Registry; PALGA, Automated National Pathological Anatomy Archive; SOTR, solid organ transplant recipient.

The primary outcome was the diagnosis of a first metachronous CSCC, defined as a new primary CSCC occurring ≥3 months after the index case. While not universally established in the literature, with intervals ranging from 2 months to 2 years, ^21,31–33^ we adopted this 3-month metachronous cutoff for two methodological reasons: (1) to minimize the risk of double-counting the same tumor (e.g., a diagnostic biopsy followed by therapeutic excision), and (2) to exclude lesions likely present but undiagnosed at the time of index visit. After exclusion of recurrences as described, tumors occurring within this three-month window were classified as synchronous and present at baseline. Patients with these synchronous tumors were retained in the cohort, with one lesion randomly selected as the index CSCC (Figure 1).

Time-to-event was the interval between the index CSCC diagnosis and the first metachronous CSCC, death, or end of follow-up, whichever occurred first. To avoid model instability due to sparse late occurrences, follow-up was censored at 10 years.

### Model development, evaluation and validation

We employed the Fine and Gray regression model (FGM) to predict first metachronous CSCC, while accounting for the competing risk of death. The FGM estimates time-dependent subdistribution hazard ratios (SHR), allowing the regression coefficients to directly quantify the effect of covariates on the cumulative incidence, thereby corresponding to the absolute risk of a metachronous CSCC at a given timepoint.^34^

Predictors, specified *a priori* based on clinical expertise and data availability, were all recorded at the time of the index CSCC. They included: age at index diagnosis, sex, history of hematologic malignancy (none vs. chronic lymphocytic leukemia / small lymphocytic leukemia (CLL/SLL) or other HM), BCC history (0 vs. 1, 2, 3, 4 or 5+ BCCs), AK history (no vs. yes), presence of synchronous CSCC (no vs. yes), CSCC differentiation grade (well vs. moderately or poorly differentiated) and tumor location (trunk vs. face, scalp and neck or extremities).

Differentiation grade was extracted from pathology reports with a free-text algorithm. Tumors were considered well-differentiated if the report did not state poor or moderate differentiation grades (n= 37% of cases).

Missing data for tumor location (3%, n=383) were handled by complete case analysis (CCA) (n=11,737), as sensitivity analyses showed no improvement in model performance with multiple imputation. We modeled a linear specification for age as it offered comparable performance to restricted cubic splines but with improved interpretability. Model performance comparisons can be found in Table S2. We evaluated proportional subdistribution hazards assumptions with methods adapted for competing risks settings,^35^ with no major violations observed.

Internal validation was primarily employed with 10-fold cross-validation. Patients were randomly assigned into 10 equal-sized folds balanced by event status. For each fold *i*, the model was fitted on the remaining 9 folds, with predictions therafter generated on the held-out *i* fold to ensure unbiased evaluation on unseen data. Bootstrap validation (n=200 replications) was performed as a sensitivity analysis and showed nearly identical performance metrics (Table S3).

Model performance was assessed for discrimination and calibration. Discrimination reflects the ability to distinguish between patients who do and do not develop a metachronous CSCC, and was measured using the time-dependent concordance index (c-index) adapted for competing risks.^35^

Time-dependent calibration plots were developed to assess the agreement between predicted and observed risks of metachronous CSCC at 1, 3, 5, and 10 years of follow-up. Calibration was evaluated on the cross-validated predictions derived from the FGM. For each time point, pseudo-observations of the cumulative incidence function were computed to obtain individual-level estimates of the observed probability of the event, accounting for the competing risk of death.^36^ Smooth calibration curves with 95% confidence intervals were then generated by regressing the observed pseudo-values on the predicted risks using locally estimated scatterplot smoothing (LOESS).^35^

To investigate potential clinical utility to safely exempt low-risk patients from follow-up, we calculated the negative predictive value (NPV) for different risk thresholds at 1-, 3-, and 5-year time horizons. All analyses were performed in R version 4.5.0, with a two-sided p-value < 0.05. The final model formula and an example calculation at 3 years after initial diagnosis are provided as Supplementary Material (Supplementary text and Table S1).

## RESULTS

### Study population

The final model included 11,737 patients diagnosed with a first CSCC in 2007 or 2008 (Table 1). Median follow-up duration was 9.0 years (interquartile range [IQR]: 3.8-10). Median age at first CSCC diagnosis was 76 years (IQR: 67-83), and 57% of patients were male. History of hematologic malignancy was seen in 3.3% (n=385). Overall, 54% of patients died during follow-up (n = 6,318/11,737).

**Table 1.**
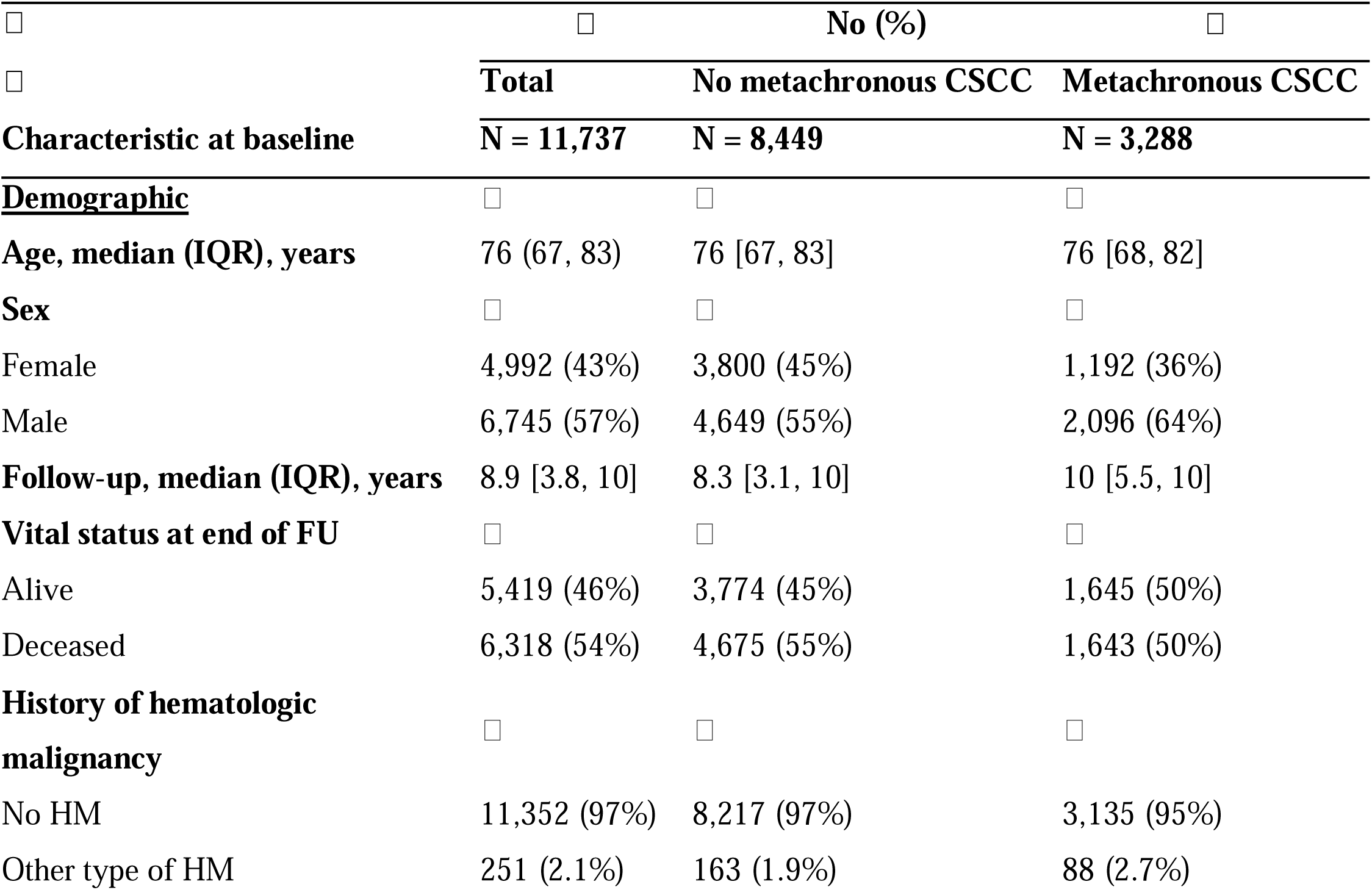
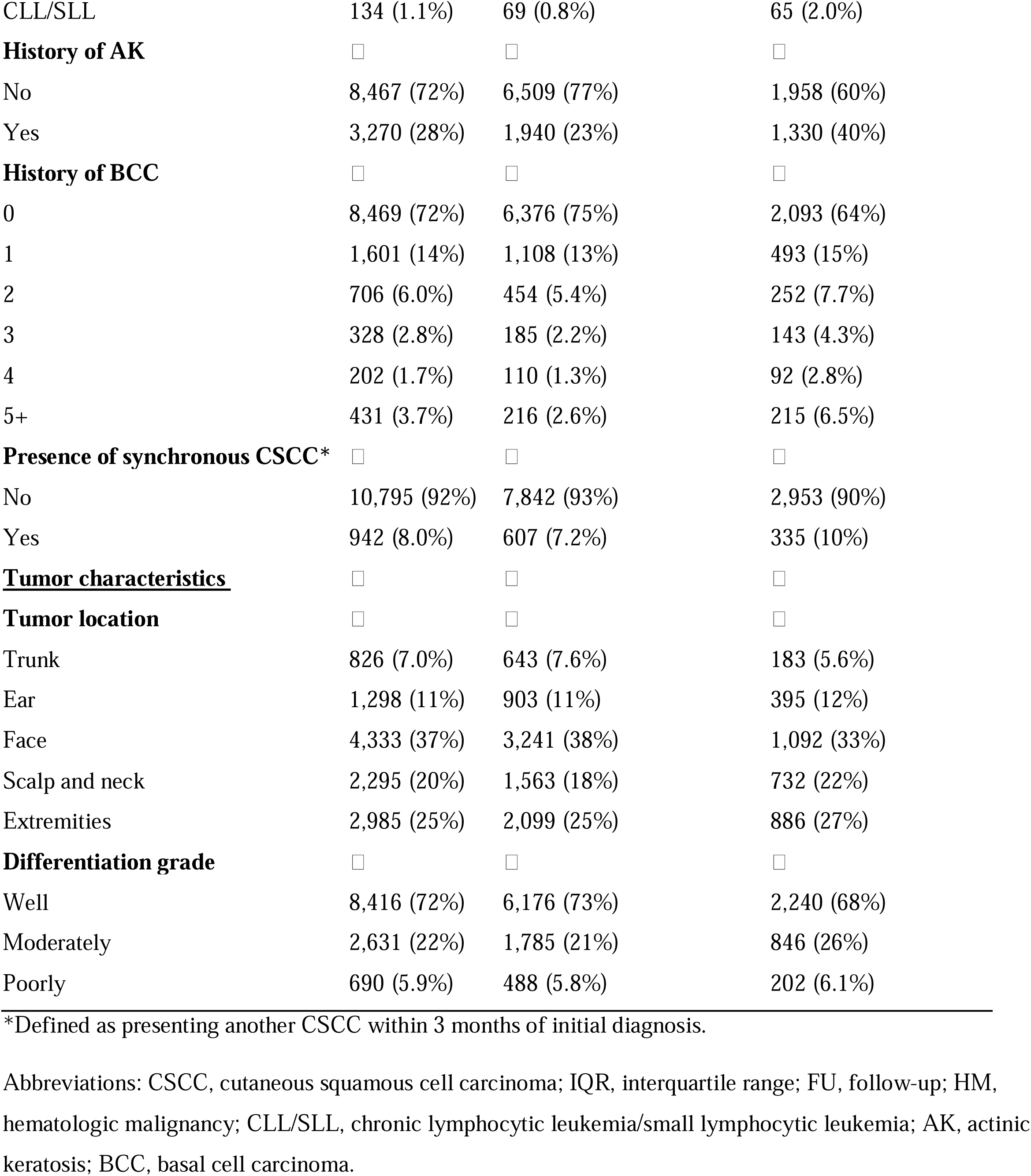
Baseline characteristics of patients with first cutaneous squamous cell carcinoma stratified by metachronous tumor development.

|  | No (%) |  |  |
| --- | --- | --- | --- |
|  | Total | No metachronous CSCC | Metachronous CSCC |
| Characteristic at baseline | N = 11,737 | N = 8,449 | N = 3,288 |
| **Demographic** |  |  |  |
| Age, median (IQR), years | 76 (67, 83) | 76 [67, 83] | 76 [68, 82] |
| Sex |  |  |  |
| Female | 4,992 (43%) | 3,800 (45%) | 1,192 (36%) |
| Male | 6,745 (57%) | 4,649 (55%) | 2,096 (64%) |
| Follow-up, median (IQR), years | 8.9 [3.8, 10] | 8.3 [3.1, 10] | 10 [5.5, 10] |
| Vital status at end of FU |  |  |  |
| Alive | 5,419 (46%) | 3,774 (45%) | 1,645 (50%) |
| Deceased | 6,318 (54%) | 4,675 (55%) | 1,643 (50%) |
| History of hematologic malignancy |  |  |  |
| No HM | 11,352 (97%) | 8,217 (97%) | 3,135 (95%) |
| Other type of HM | 251 (2.1%) | 163 (1.9%) | 88 (2.7%) |
| CLL/SLL | 134 (1.1%) | 69 (0.8%) | 65 (2.0%) |
| **History of AK** | ☐ | ☐ | ☐ |
| No | 8,467 (72%) | 6,509 (77%) | 1,958 (60%) |
| Yes | 3,270 (28%) | 1,940 (23%) | 1,330 (40%) |
| **History of BCC** | ☐ | ☐ | ☐ |
| 0 | 8,469 (72%) | 6,376 (75%) | 2,093 (64%) |
| 1 | 1,601 (14%) | 1,108 (13%) | 493 (15%) |
| 2 | 706 (6.0%) | 454 (5.4%) | 252 (7.7%) |
| 3 | 328 (2.8%) | 185 (2.2%) | 143 (4.3%) |
| 4 | 202 (1.7%) | 110 (1.3%) | 92 (2.8%) |
| 5+ | 431 (3.7%) | 216 (2.6%) | 215 (6.5%) |
| **Presence of synchronous CSCC\*** | ☐ | ☐ | ☐ |
| No | 10,795 (92%) | 7,842 (93%) | 2,953 (90%) |
| Yes | 942 (8.0%) | 607 (7.2%) | 335 (10%) |
| **<u>Tumor characteristics</u>** | ☐ | ☐ | ☐ |
| **Tumor location** | ☐ | ☐ | ☐ |
| Trunk | 826 (7.0%) | 643 (7.6%) | 183 (5.6%) |
| Ear | 1,298 (11%) | 903 (11%) | 395 (12%) |
| Face | 4,333 (37%) | 3,241 (38%) | 1,092 (33%) |
| Scalp and neck | 2,295 (20%) | 1,563 (18%) | 732 (22%) |
| Extremities | 2,985 (25%) | 2,099 (25%) | 886 (27%) |
| **Differentiation grade** | ☐ | ☐ | ☐ |
| Well | 8,416 (72%) | 6,176 (73%) | 2,240 (68%) |
| Moderately | 2,631 (22%) | 1,785 (21%) | 846 (26%) |
| Poorly | 690 (5.9%) | 488 (5.8%) | 202 (6.1%) |

\*Defined as presenting another CSCC within 3 months of initial diagnosis.
Abbreviations: CSCC, cutaneous squamous cell carcinoma; IQR, interquartile range; FU, follow-up; HM, hematologic malignancy; CLL/SLL, chronic lymphocytic leukemia/small lymphocytic leukemia; AK, actinic keratosis; BCC, basal cell carcinoma.

In total, 28% (n=3288) developed a first metachronous CSCC during follow-up within a median time of 2.5 years (IQR: 0.9-5.1). Cumulative incidence was higher (39,5%) among hematological malignancy patients. Those who developed a metachronous CSCC were more likely to be male (64% vs. 55%) and have a prior history of AK (40% vs. 23%) or BCC (36% vs. 25%) (Table 1). The most common location for the index tumor was the face (37%), followed by the extremities (25%).

### Model predictors

All pre-specified predictors were retained in the final FGM as internal validation showed no evidence of optimism (Table S3). SHR and 95% confidence intervals (CIs) are presented in Table 2. The strongest predictors of a first metachronous CSCC were history of AK (SHR 1.82, 95% CI 1.69-1.96), history of ≥ 5 BCCs (SHR 1.81, 95% CI 1.65-2.09), and history of CLL/SLL (SHR 1.78, 95% CI 1.35-2.34). Male sex, synchronous CSCC (SHR 1.28, 95% CI 1.14-1.45), moderate tumor differentiation, and primary tumor location on sun-exposed areas were also associated with higher risk. We observed no association with increasing age (SHR 0.97 per decade, 95% CI: 0.95-1.00). None of the interaction terms tested for field cancerization markers (BCC history, AK history, synchronous tumors) were statistically significant at α=0.05.

**Table 2.** Multivariable Fine and Gray regression model for the first metachronous cutaneous squamous cell carcinoma (CSCC).

| Characteristic | Numbers | Metachronous CSCC |
| --- | --- | --- |
|  |  | SHR (95% CI) |
| **Age (per 10 years increase)** | 11,737 (100) | 0.97 (0.95, 1.00) |
| **Sex** | □ | □ |
| Female | 4,992 (43%) | Ref |
| Male | 6,745 (57%) | 1.30 (1.20, 1.40)** |
| **History of hematologic malignancy** | □ | □ |
| No | 11,352 (97%) | Ref |
| Other type of HM | 251 (2.1%) | 1.25 (1.00, 1.56) |
| CLL/SLL | 134 (1.1%) | 1.78 (1.35, 2.34)** |
| **History of BCC, no. of previous diagnoses** | □ | □ |
| 0 | 8,469 (72%) | Ref |
| 1 | 1,601 (14%) | 1.18 (1.07, 1.30)* |
| 2 | 706 (6.0%) | 1.32 (1.16, 1.51)** |
| 3 | 328 (2.8%) | 1.63 (1.16, 1.51)** |
| 4 | 202 (1.7%) | 1.68 (1.36, 1.94)** |
| 5+ | 431 (3.7%) | 1.81 (1.65, 2.09)** |
| **History of AK** | □ | □ |
| No | 8,467 (72%) | Ref |
| Yes | 3,270 (28%) | 1.82 (1.69, 1.96)** |
| **Presence of synchronous CSCC** | □ | □ |
| No | 10,795 (92%) | Ref |
| Yes | 942 (8.0%) | 1.28 (1.14, 1.45)** |
| **Tumor location (first CSCC)** | □ | □ |
| Trunk | 826 (7.0%) | Ref |
| Ear | 1,298 (11%) | 1.26 (1.05, 1.50)* |
| Face, other | 4,333 (37%) | 1.11 (0.95, 1.29) |
| Scalp and neck | 2,295 (20%) | 1.32 (1.12, 1.55)** |
| Extremities | 2,985 (25%) | 1.43 (1.22, 1.67)** |
| **Differentiation grade (first CSCC)** | □ | □ |
| Well | 8,416 (72%) | Ref |
| Moderately | 2,631 (22%) | 1.25 (1.15, 1.36)** |
| Poorly | 690 (5.9%) | 1.13 (0.98, 1.31) |

\* $P$ -value < .05, \*\* < .001.
Abbreviations: CSCC, cutaneous squamous cell carcinoma; SHR, subdistribution hazard ratio; CI, confidence interval; HM, hematologic malignancy; CLL/SLL, chronic lymphocytic leukemia/small lymphocytic leukemia; AK, actinic keratosis; BCC, basal cell carcinoma.

### Model Performance

The model achieved moderate discrimination with a cross-validated c-index of 0.64 (95% CI: 0.63-0.66) at 5 years. Time-dependent calibration plots demonstrated good agreement between predicted and observed risks at 1, 3, 5, and 10 years (Figure 2). For predictions on the higher risk range (above 40%), the model tended to overestimate risk. However, further interpretation is limited by the wide confidence intervals due to sparse data in this stratum.

**Figure 2.**
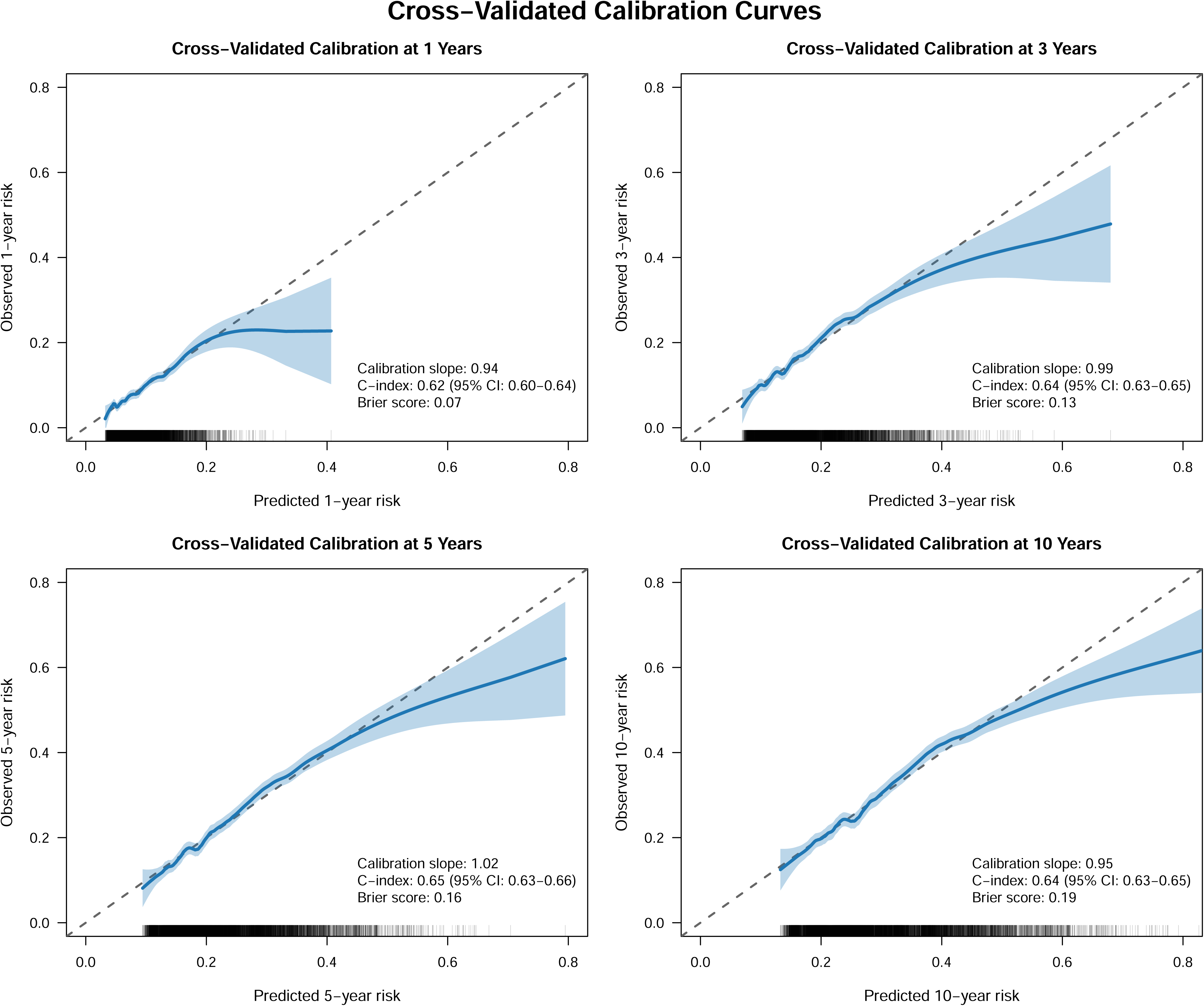
Cross-validated calibration curves for metachronous CSCC at 1, 3, 5, and 10 years. The blue line shows predicted vs observed risk with 95% CIs (LOESS smoothing). Rug plots indicate predicted risk density distribution. Abbreviations: CSCC, cutaneous squamous cell carcinoma; CI, confidence intervals.

### Clinical Utility

To assess model consistency, we compared mean predicted 5-year risk with mean observed risks across all predictor subgroups, revealing strong calibration (Figure 3). For example, the model accurately captured the stepwise risk increase with BCC count, from approximately 22% for patients with no BCC history to nearly 40% for those with five or more. Similarly, the model’s predicted 5-year risk for patients with history of AK (∼30%) aligned with the true observed risk and was substantially higher than for those without AK (∼18%).

**Figure 3.**
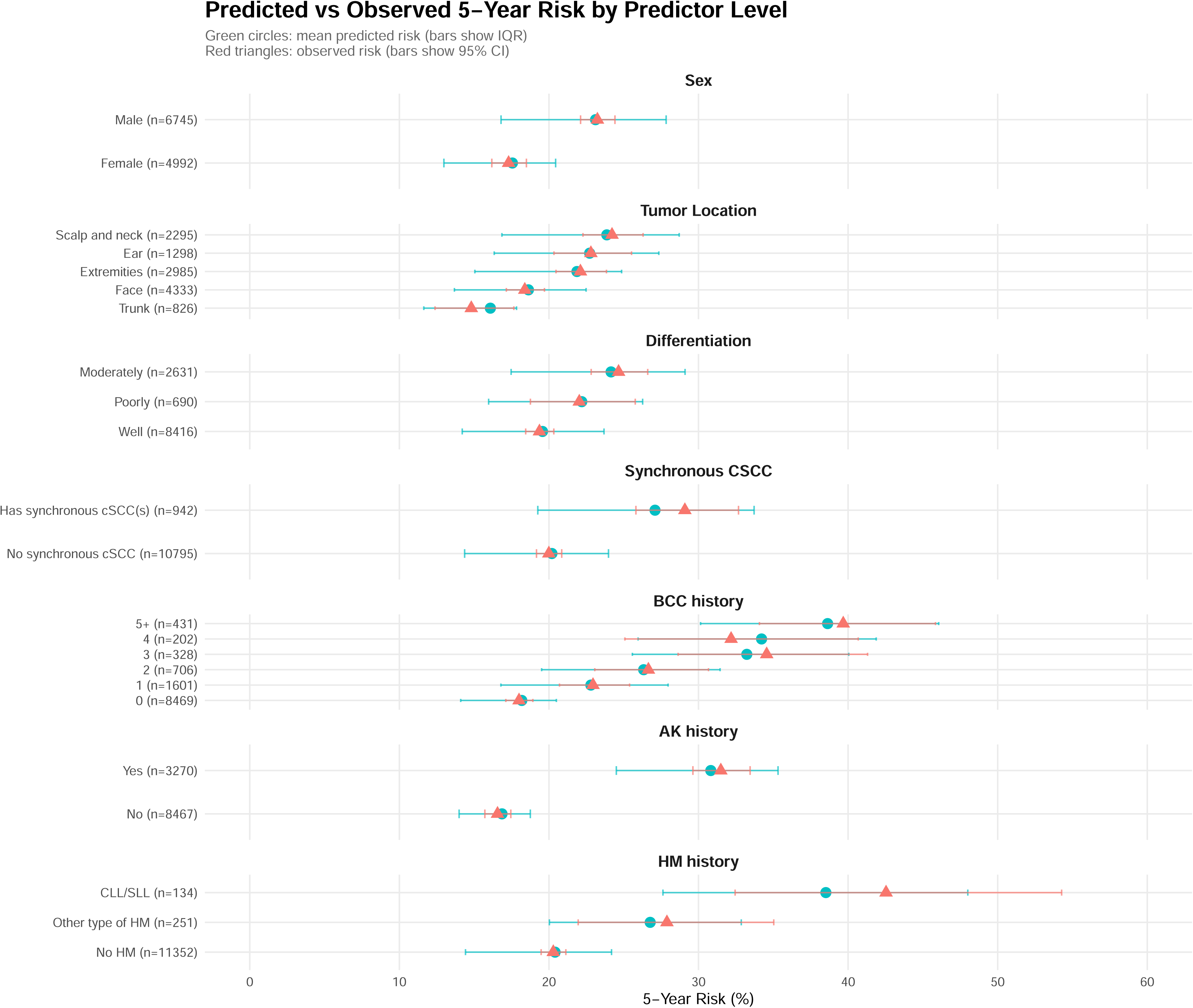
Predicted vs observed 5-year risk of metachronous CSCC by predictor level. Green circles and bars show median (IQR) predicted risks; red triangles and lines show observed risks (95% CI). Abbreviations: CSCC, cutaneous squamous cell carcinoma; CI, confidence interval; HM, hematologic malignancy; CLL/SLL, chronic lymphocytic leukemia/small lymphocytic leukemia; AK, actinic keratosis; BCC, basal cell carcinoma.

Conversely, boxplots of risk distribution showed considerable overlap between patient outcomes (Figure S1), visually confirming the model’s modest utility for precise individual risk stratification. This was quantified by a low-risk threshold analysis, where a 5-year risk threshold of 10% achieved a perfect NPV (1.00), but classified only 7 patients (<0.1%) as low-risk (Table S4).

## DISCUSSION

Correctly attributing individual risk to CSCC patients is crucial for optimizing follow-up recommendations and reducing the increasing burden of dermatological care. In this nationwide cohort study, we developed and internally validated a competing-risk adapted prognostic model for a first metachronous CSCC.

We identified clinical markers of cumulative ultraviolet (UV) exposure as robust risk indicators within our cohort. History of AK, with a nearly 2-fold increased risk, cumulative burden of pior BCCs, and primary tumor location in all sun-exposed areas were among the strongest predictors. This finding is biologically plausible due to both possible progression of AK to CSCC,^37,38^ and the broader concept of field cancerization, where cumulative UV exposure induces widespread genomic alterations in keratinocytes that predispose to multiple tumors.^39–41^ This underscores that the UV-induced (pre)cancer itself, a distinct skin aging component apart from other photodamage phenotypes (e.g., wrinkling, atrophy)^42^ may drive the risk. Remarkably, these markers remain strong indicators even within a population already diagnosed with CSCC.^3,20,43^

History of hematologic malignancy, particularly CLL/SLL, is another strong predictor in our model. The mechanism underlying this association is well-studied, reflecting both disease-related immune dysregulation and treatment-related immunosuppression, both of which impair immune surveillance against keratinocyte carcinogenesis.^44–46^ Our findings quantify this association, aligning with prior reports of more aggressive CSCC in CLL patients and further supporting clinical recommendations for enhanced surveillance and a lower biopsy threshold in this group. ^47–49^

Presence of synchronous CSCC was associated with a modest but significant 1.3-fold increased risk of a first metachronous CSCC. Although two prior studies have reported stronger associations (with HRs up to 2.5),^50,51^ those findings have limited comparability; one was a single-center study combining BCC and CSCC outcomes, and the other was a BCC-only cohort. Our nationwide CSCC-specific evidence supports adding synchronous tumors as an independent predictor, reinforcing their role in individualized risk stratification.

While some smaller studies identified age as a predictor of subsequent cSCCs,^21,23^ we did not observe this association. This likely reflects both the advanced median age at diagnosis in our cohort, attenuating its predictive effect, and the influence of survivor bias. Older patients may die from unrelated causes before developing an additional tumor, and not accounting for this can overstate the effect of age. By applying Fine–Gray competing risks regression, we provide more realistic absolute risk estimates in an elderly population.^52,53^

Our model achieved a c-index of 0.64, which, although modest, is comparable to other keratinocyte carcinoma models developed in cohorts conditioned on a first malignancy.^54–56^ For example, a CSCC cox proportional hazards model for patients with AKs reported a comparable C-index of 0.60, ^55^ and a negative binomial model for any CSCC showed good overall performance but significantly lower discrimination within the subgroup of patients with prior history of AK or CSCC (AUC ∼0.85 versus ∼0.70-0.75, respectively).^56^

This limited discrimination potentially reflects the index-event bias: conditioning on the first tumor selects a rather homogeneous and potentially high-susceptibility group, reducing variability and diminishing the predictive contribution of shared risk factors such as older age, UV skin damage and male sex.^57–59^ Moreover, the multifactorial etiology of CSCC limits precise prediction. In the negative binomial model by Wang et al., even after correcting for unmeasured interpersonal variation and incorporating genetic data, ^56^ their estimates explained only approximately 75% of the risk. This underscores that residual heterogeneity likely arises from unmeasured factors (e.g., lifetime UV exposure) or yet unknown genetic susceptibility loci.

From a clinical perspective, our model provides valuable insights into risk assessment despite moderate discrimination. The model’s excellent calibration, which allows accurately ranking patients by their risk profiles according to biologically plausible risk factors, supports its utility. Importantly, even patients with seemingly low-risk profiles (e.g., women without prior AK or BCC) still face a 5-year median risk of 15–20% for a metachronous tumor.

Strengths of this study include use of a nationwide population-based cohort with long-term follow-up, comprehensive pathology-confirmed data, and internal validation with no evidence of model overfitting. Additionally, robust methodology with competing risk regression provides a more realistic estimate of the absolute risk than traditional survival analyses and the discussed negative binomial prediction model for CSCC.

Our study also has limitations. First, registries lacked data on potentially significant predictors like Fitzpatrick skin type, lifetime UV exposure, genetic markers, and family history of skin cancer, which may have limited model performance. However, based on prior literature, we expect the inclusion of these factors would result in only moderate performance improvement. Second, the true magnitude of the AK effect is likely underestimated as our data only included histologically confirmed lesions, while most clinically diagnosed AKs are not biopsied. We recognize the challenge in distinguishing new primaries from local recurrences; however, the rule-based algorithm and the 3-month exclusion window for recurrent tumors substantially reduce its likely impact. Third, we could not find a comparable nationwide cohort with coverage of subsequent CSCCs to externally validate our model.

Finally, because our cohort included patients with multiple CSCCs, we explored statistical approaches for recurrent events (marginal Fine-Gray and Joint Frailty models) but elected to focus on the first metachronous CSCC. These methods introduce substantial computational limitations in large cohorts with competing events and significant complexity in clinical interpretation of their results.^60,61^ Also, the primary research gap lies in identifying low-risk patients for safe follow-up exemption, and our prior study has shown that the risk of new lesions escalates steeply after each new diagnosis^15^; therefore, we concentrated on predicting the first metachronous CSCC as the most clinically relevant outcome.

## CONCLUSION

This prediction model, developed and internally validated using a nationwide cohort, provides individualized absolute risk estimates for a first metachronous CSCC. It accounts for competing risk of death and uses readily available clinical and pathological factors. Despite modest discrimination, the model quantifies how established predictors collectively shape risk in this high-susceptibility population. In its current form, it provides a foundation for risk-based surveillance, supporting more precise patient counseling for high-risk profiles (e.g., those with a history of UV-induced (pre)cancer or CLL/SLL). To further enhance its clinical utility, external validation and, potentially, the identification of novel predictors are critical for refining this tool and advancing personalized dermatologic care.

## IRB approval status

This study was approved by the scientific committees of the NCR, Palga, Erasmus Medical Center (MEC-2020-0054), and Dutch Clinical Research Foundation (W20.048/NWMO20.02.007).

## Data availability

The data presented in this study are provided by the Netherlands Cancer Registry to the Erasmus MC Institute and are therefore not publicly available. However, these data may be available upon reasonable request to the corresponding author and with permission of the Netherlands Cancer Registry.

## Ethics statement and patient consent

According to the Central Committee on Research involving Human Subjects (CCMO), this observational, non-interventional study does not require approval from an ethics committee. A waiver of informed consent was granted following the code of conduct of the Foundation Federation of Dutch Medical Scientific Societies.

## CReDiT

Design: AH, CJE, BRP, MW, LH

Data collection: AH, CJE, ML

Analysis: AH, CJE, BRP

Interpretation: AH, CJE, BRP, ML, AM, TN, MW, LH

Writing: AH, CJE, BRP, AM, TN, MW, LH

## ABBREVIATIONS

CSCC: Cutaneous squamous cell carcinoma
Palga: Automated National Pathological Anatomy Archive
NCR: Netherlands Cancer Registry
ICD-O-3: International Classification of Diseases for Oncology, third edition
SOTR: Solid organ transplant recipients
NOTR: Netherlands Organ Transplant Registry
BCC: Basal cell carcinoma
AK: Actinic keratosis
HM: Hematologic malignancy
CLL/SLL: Chronic lymphocytic leukemia / Small lymphocytic leukemia
FGM: Fine and Gray model
CCA: Complete case analysis
SHR: Subdistribution hazard ratio
NPV: Negative predicted value
UV: Ultraviolet

---

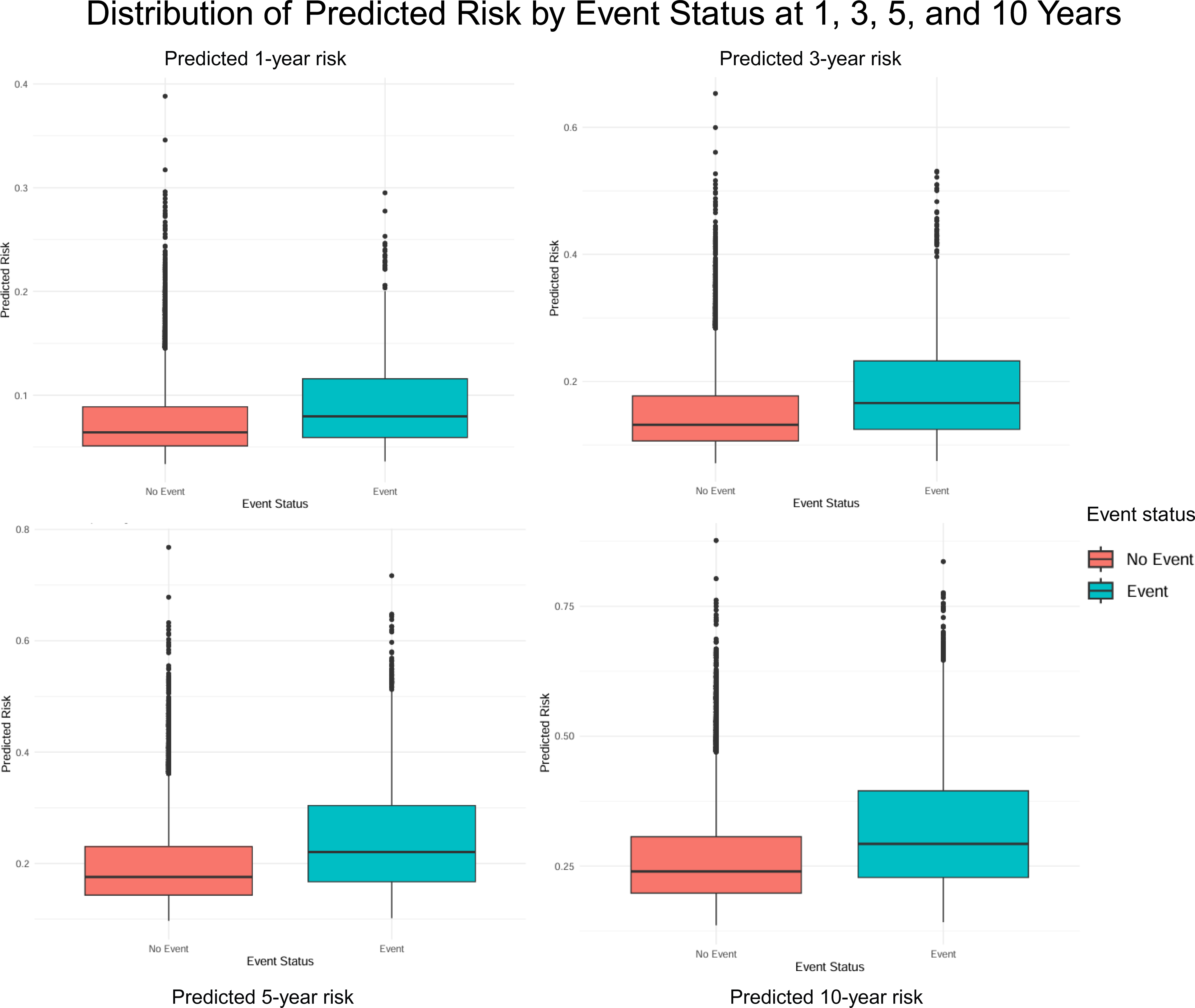

