## Supplemental material for "Predicting a metachronous cutaneous squamous cell carcinoma: a competing-risk model based on nationwide linked registries"

**Supplementary text.** Mathematical formula of the model.

**Table S1.** Calculation of the 3-year metachronous CSCC risk for an example patient**.**

**Table S2.** Model performance by specification and data handling approach.

**Table S3.** Apparent and internally validated time-dependent C-indices.

**Table S4.** NPV and coverage across risk thresholds and time horizons.

**Figure S1.** Distribution of predicted risk by event sta**tus.**

**SUPPLEMENTARY TEXT**

**MATHEMATICAL FORMULA OF THE MODEL**

$$Risk\left( t \right)= 1 - \left( 1 - Baseline_{CIF\left( t \right)} \right)^{\exp\left( LP \right)}$$

**Where:**

**Baseline_CIF(t)_** = cumulative incidence at time *t* when all predictors are at reference level

1 year = 0.0358 (3.58%)

2 years = 0.0576 (5.76%)

3 years = 0.0755 (7.55%)

5 years = 0.1026 (10.26%)

10 years = 0.1437 (14.37%)

**Linear Predictor (LP) =** sum of (coefficient × variable value)

**LP =**

- 0.0229 × age (per 10 years)

+ 0.2624 [if male]

+ 0.2209 [if HM history: Other HM]	+ 0.5771 [if HM history: CLL/SLL]

+ 0.1643 [if Prior BCC count = 1]	+ 0.2770 [if Prior BCC count = 2]

+ 0.4875 [if Prior BCC count = 3]	+ 0.5192 [if Prior BCC count = 4]

+ 0.5920 [if Prior BCC count ≥ 5]

+ 0.5971 [if AK history: Yes]

+ 0.2503 [if Synchronous cSCC: Yes]

+ 0.2285 [if Location: Ear]	+ 0.1001 [if Location: Face]

+ 0.2778 [if Location: Scalp/Neck]	+ 0.3540 [if Location: Extremities]

+ 0.2229 [if Differentiation: Moderate]	+ 0.1225 [if Differentiation: Poor]

*Coefficients presented are from the Fine-Gray subdistribution hazard model for subsequent cSCC.*

*Note: For categorical variables, enter 1 if the category applies, 0 otherwise. For age, enter actual age divided by 10. For Prior BCC count, use only the coefficient corresponding to the exact count (e.g., if patient has 2 prior BCCs, use only the coefficient 0.2770).*

**exp(LP) =** subdistribution hazard ratio relative to baseline given the calculated linear predictor

**SUPPLEMENTARY FIGURES AND TABLES**

**Table S1. Calculation of the 3-year risk of a metachronous cutaneous squamous cell carcinoma for an example patient.**

**3-year risk of subsequent cutaneous squamous cell carcinoma (cSCC) (%) =**

$$\left( 1 - \left( 1 - Baseline_{CIF\left( 3 \right)} \right)^{\exp\left( LP \right)} \right)\times100\%$$

**Step 1: define the baseline_CIF(t)_**

**Baseline_CIF(3)_ =** 0.0755 (7.55%)

**Step 2: calculate the Linear Predictor (LP)**

**Consider a 70-year-old male patient with the following characteristics:** HM history = CLL/SLL; prior BCC count = 2; AK history = yes; synchronous cSCC = no; location = face; differentiation grade = moderate.

LP = -0.0229 × 7.0 *[age]*

+ 0.2624 × 1 *[male]*

+ 0.5771 × 1 *[CLL/SLL]*

+ 0.2770 × 1 *[Prior BCC count = 2]*

+ 0.5971 × 1 *[AK history]*

+ 0.2503 × 0 *[no synchronous cSCC]*

+ 0.1001 × 1 *[face location]*

+ 0.2229 × 1 *[moderate differentiation]*

LP = -0.1603 + 0.2624 + 0.5771 + 0.2770 + 0.5971 + 0 + 0.1001 + 0.2229

**LP = 1.8763**

**Step 2: Calculate 3-year risk**

3-year risk = (1 - (1 - 0.0755)^exp(1.8763)) × 100%

3-year risk = (1 - (0.9245)^6.5227) × 100%

3-year risk = (1 - 0.6187) × 100%

**3-year risk = 38.13%**

*Interpretation: This patient has an estimated 38.13% risk of developing subsequent cutaneous squamous cell carcinoma within 3 years.*

**Table S2. Model performance by specification and data handling approach**

| Time horizon | CAA dataset, linear age **(final model)** | CCA dataset, RCS age* | Multiple imputed dataset** |
| --- | --- | --- | --- |
| Akaike information criterion (AIC) | 59968.83 | 59908.66 | 62324.64 |
| Apparent Area Under the Curve (AUC) | 0.63 | 0.63 | 0.6322  (95% CI: 0.6316 - 0.648) |

*RCS with 4-knots.

**pooled metrics according to Robin’s rules.^1^

**Table S3. Apparent and internally validated time-dependent C-indices with 95% confidence intervals (CIs).**

| Time horizon | Apparent C-index (Full Cohort) | Validated C-index (10-Fold Cross-Validation) | Validated C-index (Bootstrap, n=200 samples) |
| --- | --- | --- | --- |
| 1 year | 0.63 (0.60-0.65) | 0.62 (0.60-0.64) | 0.62 (0.60-0.64) |
| 3 years | 0.64 (0.63-0.66) | 0.64 (0.63-0.65) | 0.64 (0.63-0.66) |
| 5 years | 0.65 (0.64-0.66) | 0.64 (0.63-0.66) | 0.65 (0.64-0.66) |
| 10 years | 0.65 (0.63-0.66) | 0.64 (0.63-0.65) | 0.64 (0.63-0.66) |

**Table S4. Negative Predictive Value (NPV) and coverage across risk thresholds and time horizons**

| Time horizon (years) | Risk threshold (%) | NPV | Patients assigned to the low-risk group | | Observed events among patients in the low-risk group | |
| --- | --- | --- | --- | --- | --- | --- |
|  |  |  | n | (% of the cohort) | n | (% of the low-risk patients) |
| **1** | **-** | **1** | **0** | **0** | **0** | **0** |
|  | 4 | 0.85 | 614 | 5,1 | 111 | 18,1 |
|  | 6 | 0.81 | 3853 | 31,8 | 901 | 23,4 |
| **3** | **-** | **1** | **0** | **0** | **0** | **0** |
|  | 8 | 0.87 | 185 | 1,5 | 28 | 15,1 |
|  | 10 | 0.84 | 1365 | 11,3 | 257 | 18,8 |
| **5** | **10** | **1** | **7** | **0,1** | **0** | **0,0** |
|  | 13 | 0.84 | 1076 | 8,9 | 198 | 18,4 |

Table Note: Negative predictive value (NPV) and cohort coverage were assessed for risk-based classification at multiple time horizons. Perfect NPV values identify extremely small subgroups, whereas modestly higher thresholds markedly expand coverage but reduce NPV, illustrating the trade-off between safety and clinical applicability.

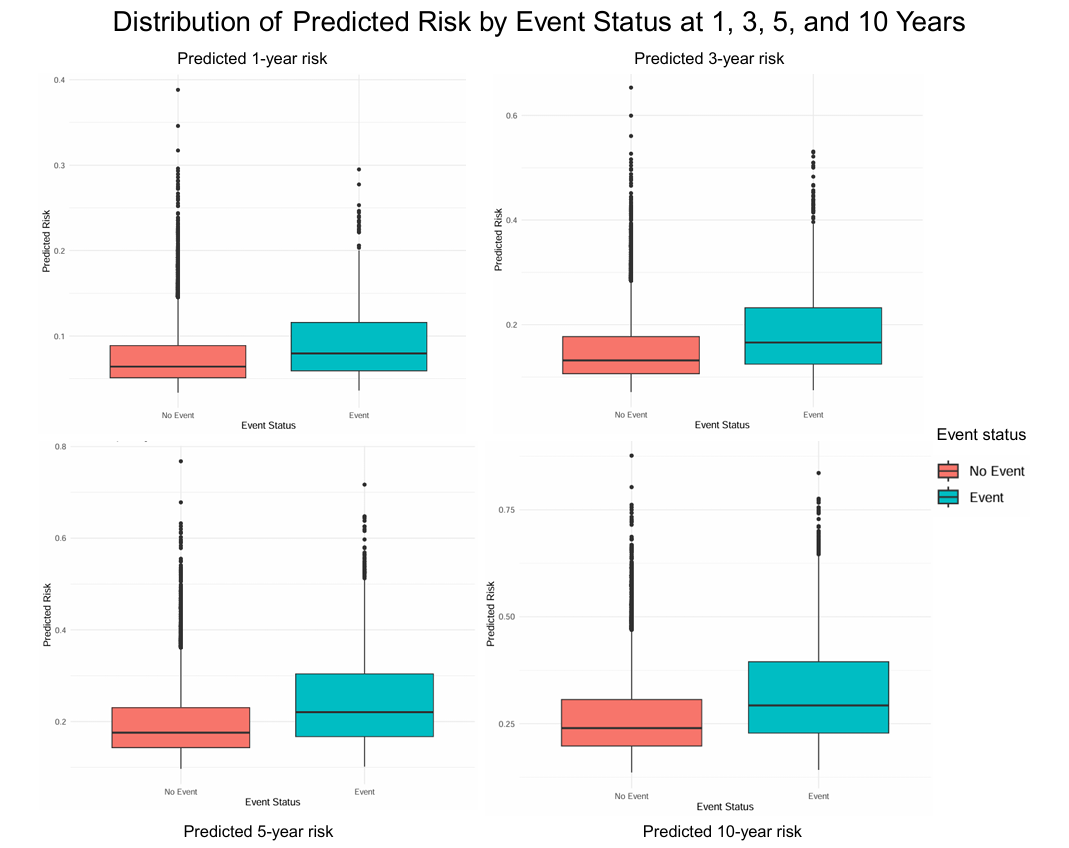

**Figure S1. Distribution of predicted risk by event status. Boxplots show predicted risks for patients with and without metachronous CSCC at 1, 3, 5, and 10 years.**

### **REFERENCES**

1. Van Buuren S. Flexible imputation of missing data: CRC press Boca Raton, FL; 2012.
